# Discovering Novel intracranial EEG Biomarkers of Seizure Generating Tissue through Time-Frequency Analysis

**DOI:** 10.64898/2026.06.12.26355482

**Authors:** Blanca Romero Milà, Nathan Phi Hoang, Marco Pinto-Orellana, Atsuro Daida, Sotaro Kanai, Naoto Kuroda, Shaun A. Hussain, Daniel W. Shrey, Eishi Asano, Hiroki Nariai, Beth A. Lopour

**Affiliations:** Department of Biomedical Engineering, University of California, Irvine, Irvine, CA, United States; Department of Pediatrics, Division of Pediatric Neurology, David Geffen School of Medicine at the University of California, Los Angeles, California, USA; Departments of Pediatrics and Neurology, Children’s Hospital of Michigan, Detroit Medical Center, Wayne State University, Detroit, MI, USA; Children’s Hospital Orange County, Division of Neurology, Orange, CA, USA; University of California, Irvine, Department of Pediatrics, Irvine, CA, USA

## Abstract

**Objective:** EEG biomarkers for seizure-generating tissue have historically been identified visually, which lacks objectivity and limits utility of automated approaches. For example, high frequency oscillations and interictal epileptiform discharges were promising markers to improve surgical outcomes for refractory epilepsy, but low specificity has hindered clinical implementation, and automated algorithms have not improved this.

**Methods:** We developed Intracranial EEG Pattern Identification and Categorization, an automated, data-driven time-frequency framework for EEG biomarker discovery. It detects transient high-power intracranial EEG waveforms (1-500 Hz) and characterizes them using eight features. In seizure-free patients, waveforms occurring predominantly in resected intracranial EEG channels are candidate biomarkers.

**Results:** In retrospective data from 14 seizure-free post-surgical patients from University of California, Los Angeles, we identified 9 waveform categories strongly associated with resected intracranial EEG channels. These included beta, gamma, and ripple band bursts, sometimes co-occurring with interictal epileptiform discharges; however, many were visually imperceptible in the broadband EEG. Using a support vector machine, we generated a unified classification metric based on these waveforms and tested it on 87 seizure-free subjects from Detroit Medical Center. This metric achieved higher area under the precision-recall curve than six state-of-the-art benchmark algorithms (p<0.001, corrected) and higher positive predictive value than three algorithms (p<0.01, corrected). Retraining the support vector machine on the Detroit dataset with five-fold cross-validation, the metric outperformed all six benchmarks across performance metrics.

**Interpretation:** Our analysis framework identified novel intracranial EEG biomarkers for seizure-generating tissue, outperforming traditional markers and generalizing across datasets, providing a new avenue for EEG biomarker discovery.

## 1. Introduction

Epilepsy is one of the most common neurological disorders, affecting between 0.5% and 1% of the population worldwide.^1^ Despite pharmacological advances, about 30% of patients remain drug-resistant.^2^ For these patients, precise localization of seizure-generating tissue is crucial, as targeted interventions such as resection, ablation, or neuromodulation can be effective. In current practice, the location of seizure onset, also called the seizure onset zone (SOZ), factors heavily into surgical planning. However, only ∼50% of patients achieve seizure freedom after resective surgery,^3^ highlighting the need for reliable biomarkers to aid in surgical decision making.

Two interictal intracranial EEG (iEEG) waveforms have been studied extensively as markers for localizing the “epileptogenic zone” (all epileptogenic tissue that, if removed, would ensure post-surgical seizure freedom). High frequency oscillations (HFOs) occur more frequently in epileptogenic than non-epileptogenic tissue across etiologies,^4–6^ and their removal correlates with favorable outcomes.^6^ However, HFOs lack sufficient reliability at the single patient level for broad clinical implementation.^7^. Moreover, although physiological and pathological HFOs likely arise from different mechanisms, their overlapping features make differentiation difficult.^8^ Interictal epileptiform discharges (IEDs) are widely used biomarkers of epilepsy,^9^ but in iEEG they are present in both seizure- and non-seizure-generating tissue, limiting specificity for surgical planning.^10^

These examples illustrate the disadvantages of relying on visual identification of EEG biomarkers: (1) It suffers from low interrater reliability^11–13^ and causes bias toward big, highly-visible events. (2) It introduces subjectivity, as definitions of EEG waveforms vary across studies. For example, HFOs are generally defined as spontaneous 80-500 Hz oscillations,^8^ but ranges vary from 30 Hz to 600 Hz,^8, 14^ and studies require anywhere from 3-6 oscillations above baseline.^8, 15, 16^ (3) It limits development of automated methods. Automated detectors for IEDs^10, 17–19^ and HFOs^20–24^ generally do not outperform visual identification for seizure localization,^25–28^ underscoring the need for more robust approaches.

Therefore, we developed intracranial EEG Pattern Identification and Categorization (i-EPIC), an objective, data-driven framework for identifying candidate biomarkers of epileptogenic tissue. Building on the s-EPIC algorithm for scalp EEG,^29^ i-EPIC uses iEEG time-frequency representations to identify waveforms that are specific to epileptogenic tissue. Time-frequency analysis can highlight oscillations at specific frequencies that might not be visible in the broadband signal, especially high frequencies which typically have a low amplitude.^30^ While time-frequency approaches have previously been used for IED^31, 32^ and HFO^30, 33–38^ detection, they typically rely on visually defined constraints for waveform features and/or fixed-duration inputs.^37, 38^ To address these limitations, i-EPIC does not make assumptions about waveform morphology or duration; it identifies iEEG patterns predominant in epileptogenic tissue and absent or rare in non-pathological regions regardless of appearance.

Here we show that i-EPIC identifies both established iEEG biomarkers (HFO- and IED-like waveforms) and novel candidates that distinguish resected from non-resected channels in seizure-free patients. We further show that waveforms identified via i-EPIC outperform existing benchmark algorithms for classification of resected channels when applied to an independent dataset.

## 2. Methods

### 2.1. Subject Information

The clinical data and iEEG for this study came from a retrospective, open-source dataset ^39^; for details, see Kuroda et al. ^40^ and Zhang et al. ^41^. All data were deidentified and publicly available, making this study exempt from IRB review.

Algorithm development utilized iEEG data from 32 pediatric patients with drug-resistant epilepsy recorded at the University of California, Los Angeles (UCLA), who were treated between August 2016 and August 2018. Of these, nine were excluded because they did not undergo resective surgery, and one due to data loading errors. Data were acquired at 2000 Hz over multiple days using subdural grid electrodes. The first nine minutes of interictal non-REM sleep data per patient were analyzed, divided into 3-minute segments

Biomarker discovery was performed using the 14 UCLA patients who achieved seizure freedom (Engel Class 1) at 24 months post-resection (13.5 [9.3 – 18] years old, 96 [69.3 – 103.5] electrodes, written as median [interquartile range (IQR)]). Seizure freedom was used as the inclusion criterion to maximize confidence that all seizure-generating tissue was resected. The remaining eight non-seizure-free subjects (13.5 [6 – 20] years old, 104 [93 – 113.5] electrodes) were reserved for an exploratory analysis.

For biomarker testing, we used 135 drug-resistant focal epilepsy patients treated at the Detroit Medical Center (DMC).^39^ Excluding adults (> 21 years old) yielded 125 subjects. Of these, 86 subjects were seizure-free post-surgery (ILAE outcome ≥ 1 years; 11 [8 – 15] years, 114 [106 – 128] electrodes) and 39 were non-seizure-free (13 [8.5 – 14] years old, 128 [112 – 131] electrodes). Surgeries were performed between January 2007 and May 2018. Recordings were acquired at 1000 Hz and consisted of 5 minutes of interictal sleep data, divided into two 2.5-minute segments for processing.

### 2.2. Biomarker discovery using intracranial EEG pattern identification and categorization (i-EPIC)

#### 2.2.1. Data pre-processing

All iEEG data underwent bipolar re-referencing (Figure 1A) and artifacts were removed using an automated envelogram-based method^42^ (Supplementary Methods S1).

**Figure 1:**
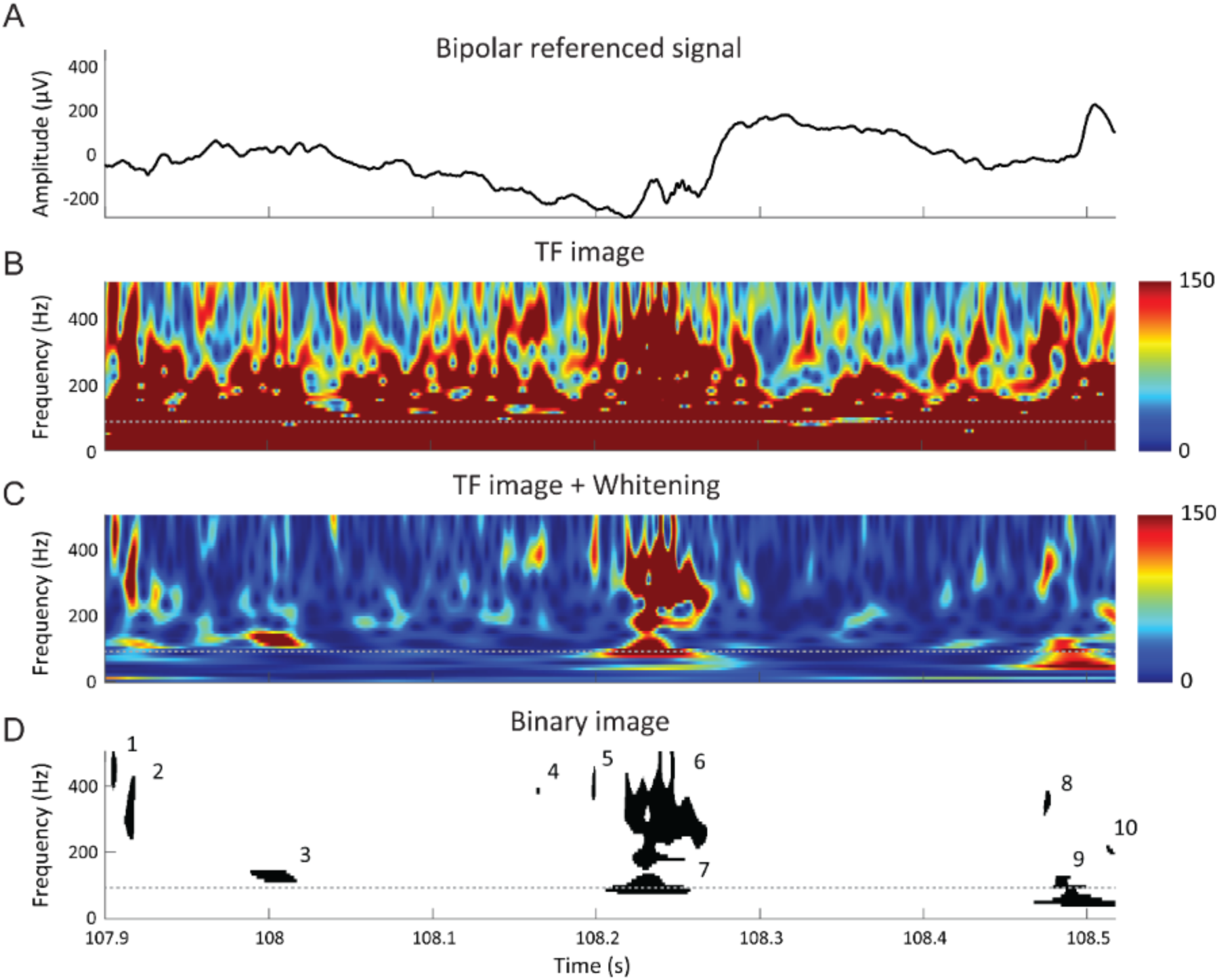
Stepwise process for extracting individual events of interest (EOIs) from iEEG data. One example is shown for a short segment of iEEG from one channel. (A) Bipolar referenced iEEG time-series, (B) time- frequency representation generated by Stockwell transform, (C) whitened time-frequency image, and (D) binarized image derived from (C), where numbered regions represent distinct detected EOIs. Note that the grey dashed line indicates the discontinuity in the time-frequency decomposition at 60 Hz.

#### 2.2.2. Time-frequency decomposition and whitening

The iEEG time-frequency decomposition was calculated using the Stockwell transform (Supplementary Methods S2) for each iEEG segment from each channel over 1-51 Hz (with a step size of 5 Hz) and 70-500 Hz (with a step of 10 Hz). This covered both low- and high-frequency bands while avoiding 60 Hz line noise (Figure 1B). Note that for the DMC dataset, the maximum frequency analyzed was 350 Hz, due to Nyquist frequency limitations. The time-frequency data were subsequently whitened using the z-score H_0_ (z_H0_) method to flatten the 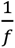 power distribution of the EEG data (Figure 1C).^43^ Then all time-frequency images were visually inspected. Channels exhibiting clear, continuous artifactual patterns were identified and excluded from further analysis.

#### 2.2.3. Identification of Events of Interest (EOIs)

For each iEEG electrode, we identified all time-frequency points that exceeded a power threshold P determined via an elbow optimization procedure (Supplementary Methods S2; Figure 1D). P was fixed across subjects, channels, and frequencies. These points were grouped into contiguous time-frequency “islands” (*_T_*_F*Is*_), individually denoted *_TFI_*_#_ (Figure 1D), and each *_TFI_*_#_ had a corresponding event of interest (EOI) in the time domain. We enforced a 10-millisecond minimum duration to capture multiple oscillations at the maximum frequency (5 oscillations at 500 Hz takes 10 ms).

#### 2.2.4. Event features

For each EOI and its associated *_TFI_*_#_, we calculated eight temporal and spectral features, adapted from Hu et al.^29^ (Figure 2). Maximum power (MP) and frequency of peak power (FPP) were extracted from the whitened TFIs. Six features were derived from the binarized image: area (A), the total time-frequency area of the event; height (H), the frequency bandwidth spanned; length (L), the temporal duration; density (D), the ratio of event area to its bounding box area; density of simultaneous events (Dsim), quantifying co-occurring activity within the event’s temporal window; and density of surrounding events (Dsur), reflecting background activity in buffer windows immediately before and after the event. To ensure consistent calculations across datasets with different sampling frequencies and frequency step sizes, pixel area in the time-frequency decomposition accounted for the frequency step size and temporal resolution at each frequency bin. For full mathematical definitions, see Supplementary Methods S3.

**Figure 2:**
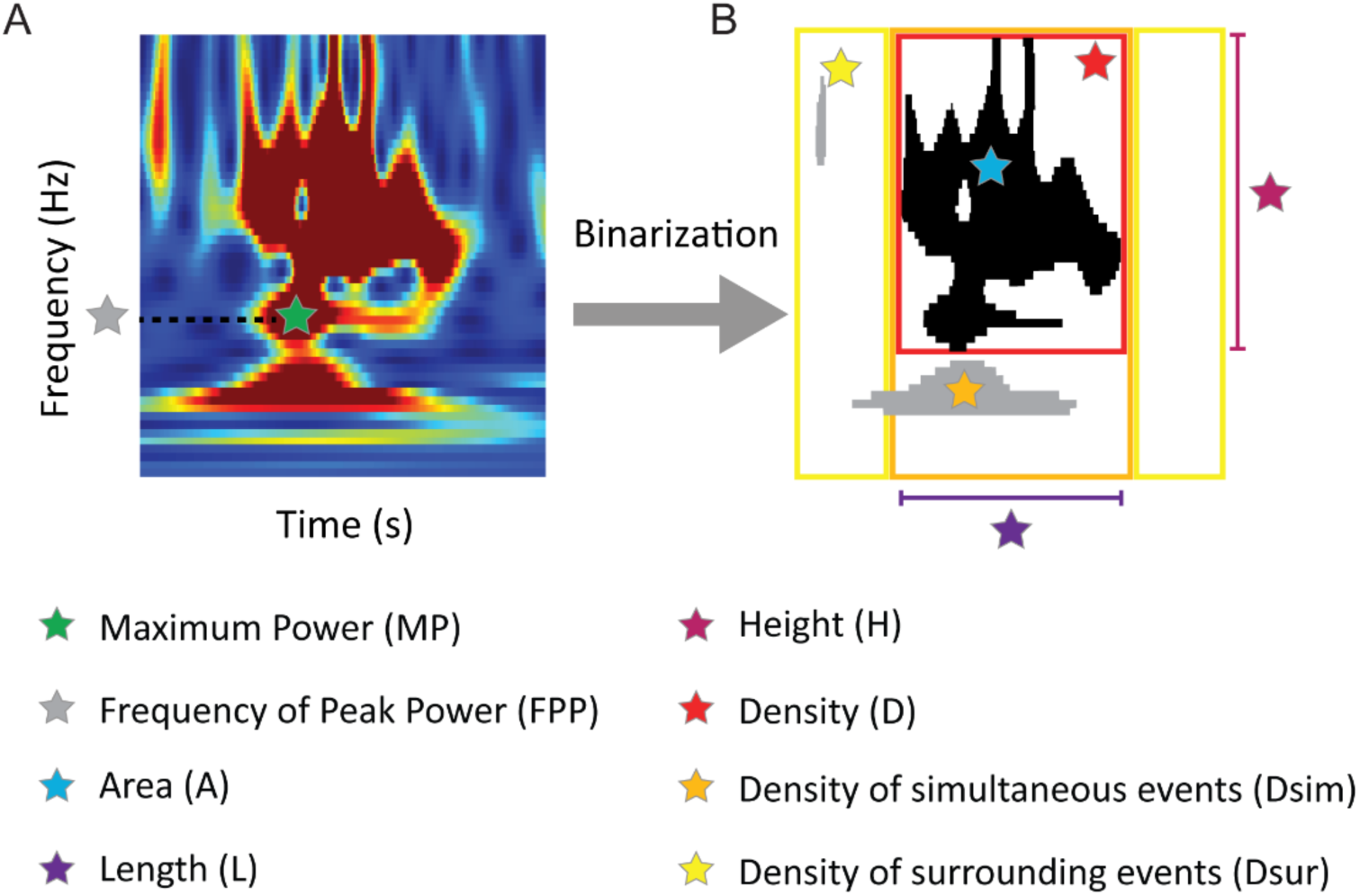
Graphic representation of EOI features. (A) The frequency of peak power (FPP, light green) and maximum power (MP, green) are shown in the whitened time-frequency decomposition. (B) The binarized time-frequency image is utilized to represent area (blue, sum of black pixels), length (purple), height (pink), density (red), density of simultaneous events (Dsim, orange), and density of surrounding events (Dsur, yellow).

#### 2.2.5. Feature Categorization

We categorized each EOI based on its combination of features. To start, each feature was discretized as follows:

*FPP* was given a value of 1 through 4, representing the frequency bands 1-51 Hz, 70-210 Hz, 211-350 Hz, and 351-500 Hz, respectively. This accounted for intrinsic differences between events with different frequencies, which are particularly pronounced when comparing the lowest and highest frequency bands.

For the features *MP, A, H, L, D*, *Dsim,* and *Dsur*, median values were calculated for each frequency band, based on events from all subjects and channels. Then features above their respective medians were assigned a value of one, while those below were assigned zero.

Consequently, each EOI was placed into a category defined by an eight-element code comprising one quaternary element (*FPP*), and seven binary elements (*MP, A, H, L, D*, *Dsim,* and *Dsur*).

#### 2.2.6. Extraction of significant categories

Categories representing candidate biomarkers satisfied three requirements. First, they must have > 500 EOIs to ensure generalizability. Second, the proportion of EOIs from resected channels must exceed the 99^th^ percentile of a binomial distribution, indicating preferential occurrence in “pathological” channels (resected channels of seizure-free patients). Any bipolar channel including at least one resected electrode was considered resected. Third, they must achieve a median AUROC across subjects >0.65 (above chance levels^44^) to ensure robust predictive power.

#### 2.2.7 Combination of significant categories using support vector machine

To integrate multiple significant EOI categories, we used a support vector machine (SVM) classifier with a radial basis function kernel. The input features for the SVM were the EOI counts in each channel for every significant category, divided by the maximum number of events across all categories and channels within each patient. The output was a binary classification predicting whether the channel was resected or non-resected. To optimize the SVM’s performance, we conducted a grid search over a range of hyperparameters, specifically the box constraint and kernel scale. The best hyperparameters were selected based on the lowest 5-fold cross-validation loss. We then trained the SVM using these optimized parameters and evaluated its performance using cross-validation.

### 2.3. Biomarker discovery through application of i-EPIC to data from UCLA

We performed the i-EPIC biomarker discovery process (Sections 2.2.1 - 2.2.4) by detecting EOIs in seizure-free patients from the UCLA dataset. The power threshold was determined using data from all seizure-free subjects and channels. To enhance generalizability, we implemented a leave-one-out cross-validation framework for all subsequent steps. Median values for feature categorization were derived from training subjects and applied to both training and test data (Section 2.2.5). Then, significant categories were identified from training data (Section 2.2.6) and normalized EOI counts per category were used to fit an SVM classifier to the training subjects to predict resected versus non-resected channel status. The model was evaluated on the held-out test subject (Section 2.2.7).

To test on the DMC data, we trained a final model using all UCLA seizure-free subjects without cross-validation. Feature medians for categorization were calculated using data from all UCLA seizure-free subjects, and EOIs were defined and categorized. Significant categories were re-defined, and the SVM classifier was trained on all subjects to generate the final predictive model.

### 2.4 Testing of iEEG biomarkers derived via i-EPIC using data from DMC

To validate the clinical relevance of the iEEG biomarkers suggested by i-EPIC, we tested the classification accuracy on an independent dataset of seizure-free DMC patients. DMC iEEG data were processed as described in Sections 2.2.1-2.2.4, using the power thresholds and feature categorization medians derived from the UCLA data (Sections 2.2.5, 2.2.6). The classification of resected and non-resected channels was tested in the DMC dataset using the final SVM model derived from the UCLA data. For DMC, the EOI counts were normalized by the maximum number of events across categories and channels for each subject. No parameters were tuned for DMC: the power threshold *_P_*, feature categorization medians, EOI categories, and SVM model were all fixed from the UCLA dataset. The only adaptation was the exclusion of the highest EOI frequency band (351-500 Hz), due to the lower sampling rate of the DMC dataset (1000 Hz).

Because the relative proportions of EOI categories may depend on the underlying etiologies of the patients and the locations of implanted electrodes, we did an exploratory analysis to measure the impact of these institution-specific factors. Following the procedure above, a new SVM classifier was trained and evaluated via 5-fold cross-validation on the DMC subjects to predict resected or non-resected channel status. In this test, the candidate biomarkers (EOI categories) were based solely on the UCLA training data, but the relative proportions of EOI categories in the SVM were tailored to DMC.

### 2.5 Comparison of i-EPIC biomarkers to state-of-the-art biomarkers of epileptogenicity

IEDs and HFOs are established biomarkers of epileptogenic tissue^45, 46^ and were thus used as benchmarks to contextualize i-EPIC biomarker performance. IEDs were automatically detected using spike morphology based on upslope, instantaneous energy, and downslope^17^, spike ripples with *PyHFO*^47^, and HFOs using four published algorithms: a detector based on the root-mean-square (RMS) amplitude^20^ with false HFO rejection,^48^ the iterative gamma-fit HFO detector (IGHD),^21^ the Hilbert detector (HIL),^22^ and the MNI detector.^23^ Data were preprocessed as in Section 2.2.1 and the classification metric was event rate per minute per channel. See full details in Supplementary Methods S4.

### 2.6. Visual inspection

To interpret the events identified by i-EPIC, three board-certified epileptologists visually reviewed a subset of representative EOIs. From each category, we selected the twenty EOIs closest to the median in the feature space. This approach ensured that reviewed events were typical examples of each category rather than outliers. In each group of 20 EOIs, we ensured that at least 50% of subjects contributed at least one EOI to assess inter-subject variability.

For each selected EOI, epileptologists were shown bipolar iEEG traces with surrounding multi-channel context and category-specific band-pass-filtered signals for review. To replicate a standard clinical scenario, reviewers were blinded to the TF representations. They were also blinded to resected volume and surgical outcome.

### 2.7. Statistical Analysis

Classification accuracy was evaluated using three metrics applied to SVM predictions for each channel. The Area Under the Receiver Operating Characteristic Curve (AUROC) provides robust overall performance measurement but can be optimistic under class imbalance, while the Area Under the Precision Recall Curve (AUPRC) is sensitive to class imbalance and scales performance relative to target class size.^49^ This is important because non-resected channels typically outnumber resected channels. Positive Predictive Value (PPV) was computed using Tukey’s upper fence to identify outlier channels as resection candidates – arguably the most clinically relevant metric for surgical planning, as it accommodates cases where no channels exceed the outlier threshold, which may occur when implanted electrodes do not directly capture seizure onset.

## 3. Results

### 3.1. i-EPIC identifies categories of EOIs that have a strong association with resected iEEG channels

Before EOI detection, iEEG time-frequency plots were inspected for artifactual channels. We excluded 35 out of 1275 channels (2.7%); see Supplementary Figure 1 for examples. Automatic artifact detection (Section 2.2.1) removed 24.6 minutes of iEEG (3.2%). A power threshold of P = 120 was determined using the elbow method, binning together time-frequency data from UCLA seizure-free subjects.

The i-EPIC pipeline (Sections 2.2.1-2.2.3) identified 929,325 EOIs across all seizure-free UCLA subjects (median: 64,551 EOIs per subject; IQR: 46,719 – 79,881, range: 16,157 – 151,632). Each EOI was characterized using eight features (Section 2.2.4).

To improve robustness, leave-one-subject-out cross-validation was implemented. Of the 340 possible EOI categories (Section 2.2.5), 96.1 [94.3 – 101] (median [IQR]) occurred significantly more frequently in resected than non-resected channels across subjects. Following Section 2.2.6, we then identified 15 categories (9 [8 – 10] across iterations) as potential markers of resected channels (Table 1). Interestingly, all 15 categories contained EOIs with FPP in the 1-51 Hz or 70-210 Hz frequency range, and 9/15 appeared in at least half of the leave-one-out iterations, demonstrating consistent patterns across the cohort.

**Table 1:** Unique EOI categories with high discriminatory power between resected and non-resected channels identified using the 14 UCLA seizure-free subjects and leave-one-out cross-validation. Each row represents the one category, and columns indicate the features used for categorization – FPP: frequency of peak power; MP: maximum power; Area; Length; Height; Density; Dsim: density of simultaneous events; Dsur: density of surrounding events. The table reports the median [interquartile range], as well as a categorical label (“high” or “low”). For all features except FFP, “high” indicates that the feature is above or equal to the median for that category, while “low” indicates it is below the median. The last column indicates the number of leave-one-out iterations in which the category was significant.

|  | FPP | MP | Area | Length | Height | Density | Dsim | Dsur | Iter. |
| --- | --- | --- | --- | --- | --- | --- | --- | --- | --- |
| 1 | 1-51<br>31 [25] | high<br>283.2 [219.2] | high<br>1.53 [1.80] | high<br>0.12 [0.12] | high<br>20 [25] | low<br>0.57 [0.24] | high<br>0.0099 [0.028] | high<br>0.0099 [0.017] | 14 |
| 2 | 1-51<br>46 [10] | high<br>206.03 [58.37] | high<br>0.65 [0.30] | low<br>0.051 [0.012] | high<br>20 [15] | low<br>0.68 [0.21] | high<br>0.010 [0.033] | high<br>0.012 [0.021] | 14 |
| 3 | 1-51<br>41 [10] | high<br>199.56 [51.52] | high<br>0.64 [0.31] | low<br>0.051 [0.013] | high<br>20 [10] | low<br>0.70 [0.18] | high<br>0 [0.0036] | low<br>0 [0.00056] | 14 |
| 4 | 1-51<br>26 [20] | high<br>281.10 [239.07] | high<br>1.69 [2.02] | high<br>0.13 [0.16] | high<br>15 [25] | low<br>0.58 [0.22] | high<br>0.00037 [0.0078] | low<br>0 [0.00056] | 14 |
| 5 | 1-51<br>51 [10] | high<br>173.84 [19.80] | low<br>0.37 [0.11] | low<br>0.039 [0.013] | high<br>10 [5] | low<br>0.76 [0.20] | high<br>0.0069 [0.028] | high<br>0.011 [0.020] | 14 |
| 6 | 1-51<br>41 [10] | low<br>143.22 [16.14] | low<br>0.25 [0.14] | low<br>0.029 [0.016] | high<br>10 [0] | low<br>0.73 [0.18] | high<br>0.0063 [0.026] | high<br>0.0099 [0.017] | 14 |
| 7 | 70-210<br>100 [40] | high<br>345.60 [515.26] | high<br>1.96 [3.21] | high<br>0.048 [0.055] | high<br>90 [100] | low<br>0.44 [0.21] | high<br>0.0021 [0.017] | low<br>0.0028 [0.0062] | 10 |
| 8 | 1-51<br>26 [30] | low<br>130.73 [12.95] | low<br>0.14 [0.11] | low<br>0.028 [0.022] | high<br>5 [0] | high<br>1 [0] | high<br>0.0026 [0.025] | high<br>0.010 [0.010] | 9 |
| 9 | 70-210<br>90 [30] | low<br>149.33 [21.82] | low<br>0.27 [0.16] | low<br>0.015 [0.0045] | low<br>20 [10] | high<br>0.81 [0.23] | high<br>0.0078 [0.031] | high<br>0.023 [0.026] | 7 |
| 10 | 1-51<br>21 [30] | low<br>130.37 [12.61] | low<br>0.14 [0.12] | low<br>0.028 [0.023] | high<br>5 [0] | high<br>1 [0] | high<br>0 [0.012] | high<br>0.0075 [0.011] | 5 |
| 11 | 1-51<br>11 [10] | low<br>144.47 [15.68] | low<br>0.36 [0.07] | high<br>0.073 [0.014] | high<br>5 [0] | high<br>1 [0] | high<br>0.00051 [0.010] | high<br>0.0063 [0.0081] | 4 |
| 12 | 1-51<br>11 [10] | low<br>145.28 [15.59] | low<br>0.36 [0.07] | high<br>0.073 [0.014] | high<br>5 [0] | high<br>1 [0] | high<br>0.0059 [0.029] | high<br>0.0088 [0.018] | 3 |
| 13 | 70-210<br>100 [30] | low<br>145.59 [16.14] | low<br>0.26 [0.13] | low<br>0.015 [0.0045] | low<br>30 [15] | low<br>0.59 [0.074] | high<br>0.010 [0.033] | high<br>0.024 [0.027] | 2 |
| 14 | 70-210<br>140 [40] | low<br>174.99 [15.22] | low<br>0.46 [0.10] | low<br>0.013 [0.003] | high<br>50 [10] | high<br>0.71 [0.073] | high<br>0.011 [0.042] | high<br>0.028 [0.036] | 1 |
| 15 | 70-210<br>80 [30] | low<br>162.23 [21.16] | low<br>0.44 [0.15] | high<br>0.026 [0.007] | low<br>30 [10] | low<br>0.57 [0.099] | high<br>0.0084 [0.029] | high<br>0.023 [0.024] | 1 |

### 3.2. No single EOI category generalizes across subjects, motivating integration with SVM

The accuracy of individual EOI categories for classifying resected and non-resected channels was assessed using leave-one-out validation (Figure 3A). Based on the AUROC values for all subjects, no single category consistently outperformed others. Six EOI categories were the most accurate for at least one subject (Figure 3B). Category 4 had the highest AUROC for the most subjects (4 out of 14 subjects). Given this inter-subject variability, we integrated the information across categories using an SVM classifier, yielding a median AUROC of 0.66 [0.61 – 0.82] (median [IQR]), AUPRC of 0.67 [0.34 - 0.82], and PPV of 1 [0.4 – 1] across seizure free UCLA subjects. Notably, although no individual category consistently outperformed the SVM, several categories achieved comparable performance (Figure 3A, Supplementary Figure 2A-B).

**Figure 3:**
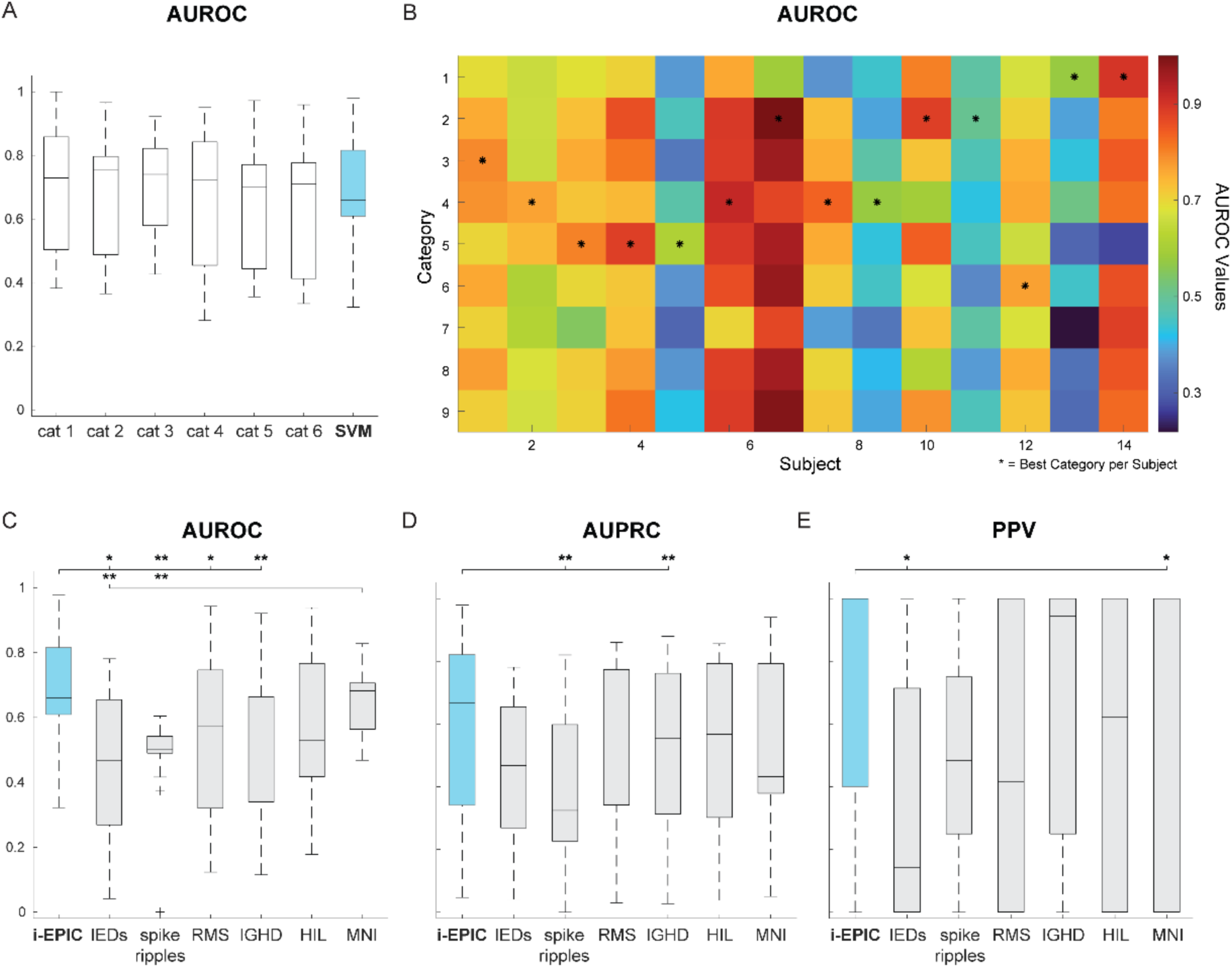
I-EPIC demonstrated superior or comparable performance compared to established SOZ biomarkers in classifying resected and non-resected channels from 14 seizure-free subjects in the UCLA dataset. (A) In the training data, no single EOI category generalizes across all subjects. Boxplots illustrate the distribution of Area Under the Receiver Operating curve (AUROC) for the six EOI categories present in all subjects, compared to the SVM. (B) AUROC values for each EOI category across individual subjects. (C) Comparison of classification accuracy for i-EPIC vs. six established detectors based on AUROC, (D) Area Under the Precision-Recall Curve (AUPRC), and (E) Positive Predictive Value (PPV). Boxplots show i-EPIC, Interictal Epileptiform Discharges (IEDs), spike ripples, root-mean-square (RMS), iterative gamma-fit HFO detector (IGHD), Hilbert detector (HIL), and MNI detectors. Central lines represent medians, boxes show interquartile ranges, and whiskers extend to the most extreme non-outlier data points. Statistical significance determined by Wilcoxon signed rank test and BH adjustment (*: p<0.05; **: p<0.01).

### 3.3. i-EPIC biomarker outperforms established SOZ biomarkers

The i-EPIC SVM biomarker was then benchmarked against established detectors for spikes, spike ripples, and HFOs (Section 2.5). Note that i-EPIC biomarkers were derived using leave-one-out cross-validation to prevent overfitting, whereas benchmark algorithms required no training and were evaluated on the full dataset.

i-EPIC significantly outperformed several of the benchmark algorithms across all three metrics (Wilcoxon signed rank test, Benjamini-Hochberg (BH) adjustment): AUROC exceeded that of IEDs, spike ripples, RMS, and IGHD detectors (Figure 3C;*: p<0.05; **: p<0.01); AUPRC exceeded spike ripples and IGHD detections (Figure 3D; *: p<0.05); and PPV exceeded IEDs and the MNI detector (Figure 3E; *: p<0.05). No benchmark detector significantly outperformed the i-EPIC SVM on any metric.

### 3.4 I-EPIC identifies candidate biomarkers in the beta, gamma, and ripple frequency bands

Visual review (Section 2.6) revealed that most events detected by i-EPIC were not discernable in the iEEG time series but were clearly visible in the TF representations (Figure 4). Categories 1-4 predominantly consisted of gamma oscillations, sometimes visible in the time series and sometimes co-occurring with spikes. Categories 5 and 6 contained smaller gamma bursts detectable only in the TF domain. Categories 8, 10, 11, and 12 featured low-amplitude beta activity, observable only in the TF domain. Category 7 included high MP and high area ripple-like events visible in both the TF images and the time series, while category 9 contained smaller events in the same frequency band, with low MP and low area. Inter-reviewer interpretation of these events varied substantially; however, a common sentiment, particularly for categories 5-9, was that many events were either not discernable in the time series or not clinically reportable.

**Figure 4:**
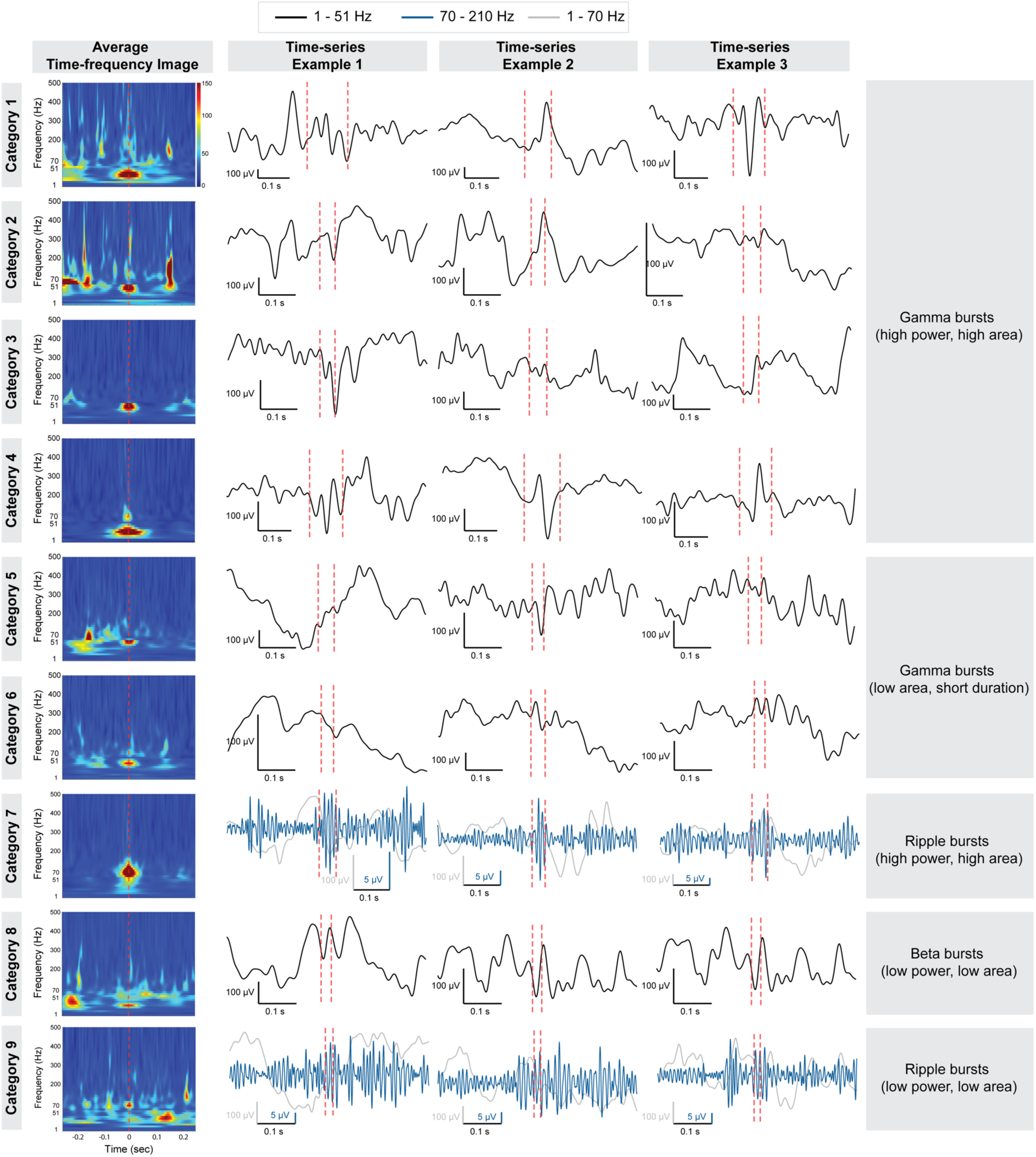
Interpretation of significant EOI categories from i-EPIC applied to UCLA seizure-free patients. For each category, the first column shows the average TF decomposition computed across the 20 representative events shown to the epileptologists for review, aligned to the event center. The three remaining columns display time-series examples from three representative events per category. Time-series signals were band-pass filtered according to category definition (1 – 51 Hz or 70 – 210 Hz, black and blue respectively). For high-frequency categories (70 – 210 Hz), the broadband 1 – 70 Hz signal (grey) is shown to provide context.

### 3.5. Testing on an independent dataset confirms i-EPIC’s superior classification capability

The i-EPIC SVM was then tested on 87 seizure-free DMC subjects as follows. A final model was trained on all 14 UCLA seizure-free subjects without cross-validation, yielding 9 categories as potential biomarkers of resected channels – the same 9 identified in at least half of the cross-validation iterations (Section 3.1, Table 1). This model was applied to the DMC dataset using UCLA-derived power thresholds, feature categorization medians, and the 9 EOI categories.

In the DMC dataset, the SVM showed significantly higher AUROC than three of the six EOI categories (Figure 5A, Wilcoxon signed rank test, BH adjustment; ***: p < 0.001; Supplementary Figure 2C-D). Performance varied substantially across subjects, with category 6 achieving the highest AUROC most frequently (25 out of 87 subjects, Figure 5B).

**Figure 5:**
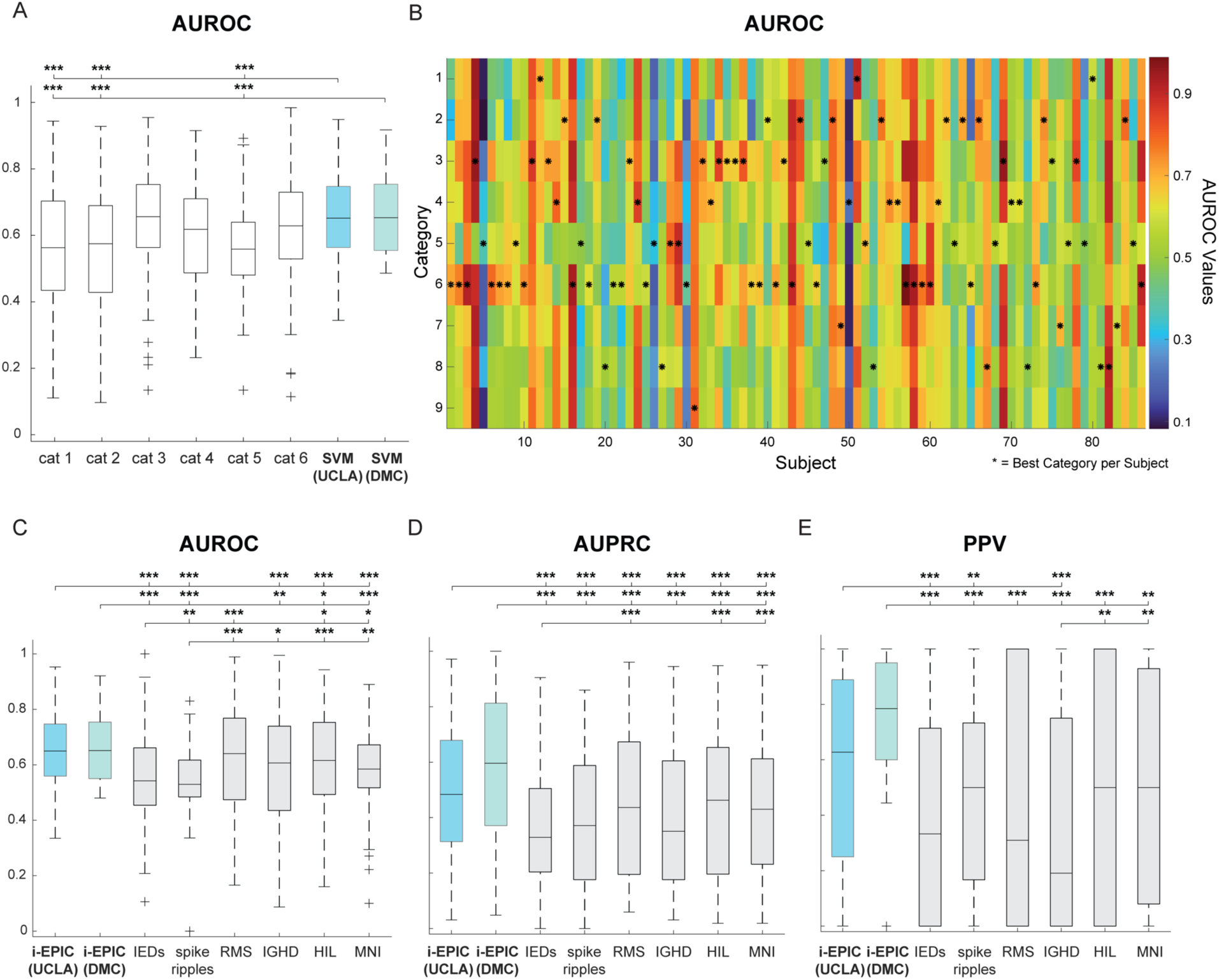
I-EPIC maintains superior performance in independent testing, outperforming established biomarkers in classifying resected and non-resected channels. (A) No single EOI category generalizes across all subjects. Boxplots illustrate the distribution of Area Under the Receiver Operating curve (AUROC) for the six EOI categories present in all subjects compared to the SVM. (B) AUROC values for each EOI category across individual subjects. (C) Comparison of classification accuracy for i-EPIC vs. six established detectors based on AUROC, (D) Area Under the Precision-Recall Curve (AUPRC), and (E) Positive Predictive Value (PPV). Data are shown for i-EPIC, Interictal Epileptiform Discharges (IEDs), spike ripples, root-mean-square (RMS), iterative gamma-fit HFO detector (IGHD), Hilbert detector (HIL), and MNI detectors. Central lines represent medians, boxes show interquartile ranges, and whiskers extend to the most extreme non-outlier data points. Statistical significance determined by Wilcoxon signed rank test and BH adjustment (*: p<0.05; **: p<0.01; ***: p<0.001).

Compared to established algorithms, i-EPIC significantly outperformed IEDs, spike ripples, IGHD, HIL, and MNI in AUROC (Figure 5C; ***: p < 0.001), all established biomarkers in AUPRC (Figure 5D; ***: p < 0.001), and IEDs, spike ripples, and IGHD in PPV (Figure 5E; **: p < 0.01, ***: p < 0.001).

To assess whether institution-specific training could improve performance, we re-trained the SVM using 5-fold cross-validation on DMC seizure-free subjects, while retaining UCLA-derived thresholds, categorization medians, and EOI categories. This model significantly outperformed all benchmark detectors based on AUROC, AUPRC, and PPV (Figure 5C, Wilcoxon signed rank test, BH adjustment; *: p<0.05, **: p < 0.01, ***: p < 0.001), except the RMS detector AUROC. This suggests that, while i-EPIC’s EOI detection and categories generalize across institutions, institution-specific calibration enhances classification performance.

### 3.6 The biomarker discovery process yields similar EOI categories for multiple training datasets

To assess the impact of the training dataset on selection of EOIs and categories, we repeated the biomarker discovery procedure (Section 2.2) using 87 seizure-free DMC subjects, with all power thresholds, categorization medians, and EOI categories derived from DMC. The resulting top-performing EOI categories showed substantial overlap with those from the UCLA dataset, with 66.67% of the 9 best-performing categories being identical across both datasets (Supplementary Table 1). The remaining categories, while not exact matches, exhibited strong similarities to their UCLA-derived counterparts.

### 3.7. i-EPIC biomarkers show promise for discrimination of seizure-free versus non-seizure-free patients

We then applied the i-EPIC algorithm to non-seizure-free patients from both datasets. This classification task is intrinsically more difficult, as both resected and non-resected channels may include both epileptogenic and non-epileptogenic tissue. Theoretically, an ideal biomarker would perform better in seizure-free compared to non-seizure-free subjects.

In the UCLA dataset, the i-EPIC SVM (Section 3.3) demonstrated significantly better PPV and AUPRC in seizure-free (n = 14) versus non-seizure-free (n = 8) subjects (Figure 6A, Supplementary Figure 4A, Wilcoxon rank-sum test, BH correction). For the DMC dataset (n = 126), the UCLA-trained SVM showed no significant difference between seizure-free (n = 87) and non-seizure-free (n = 39) subjects (Figure 6B, Supplementary Figures 3B, 4B). The DMC-retrained SVM showed a borderline difference in PPV (p = 0.048, Wilcoxon rank-sum test) that did not survive multiple-comparisons correction (Figure 6C; Supplementary Figures 3C, 4C).

**Figure 6:**
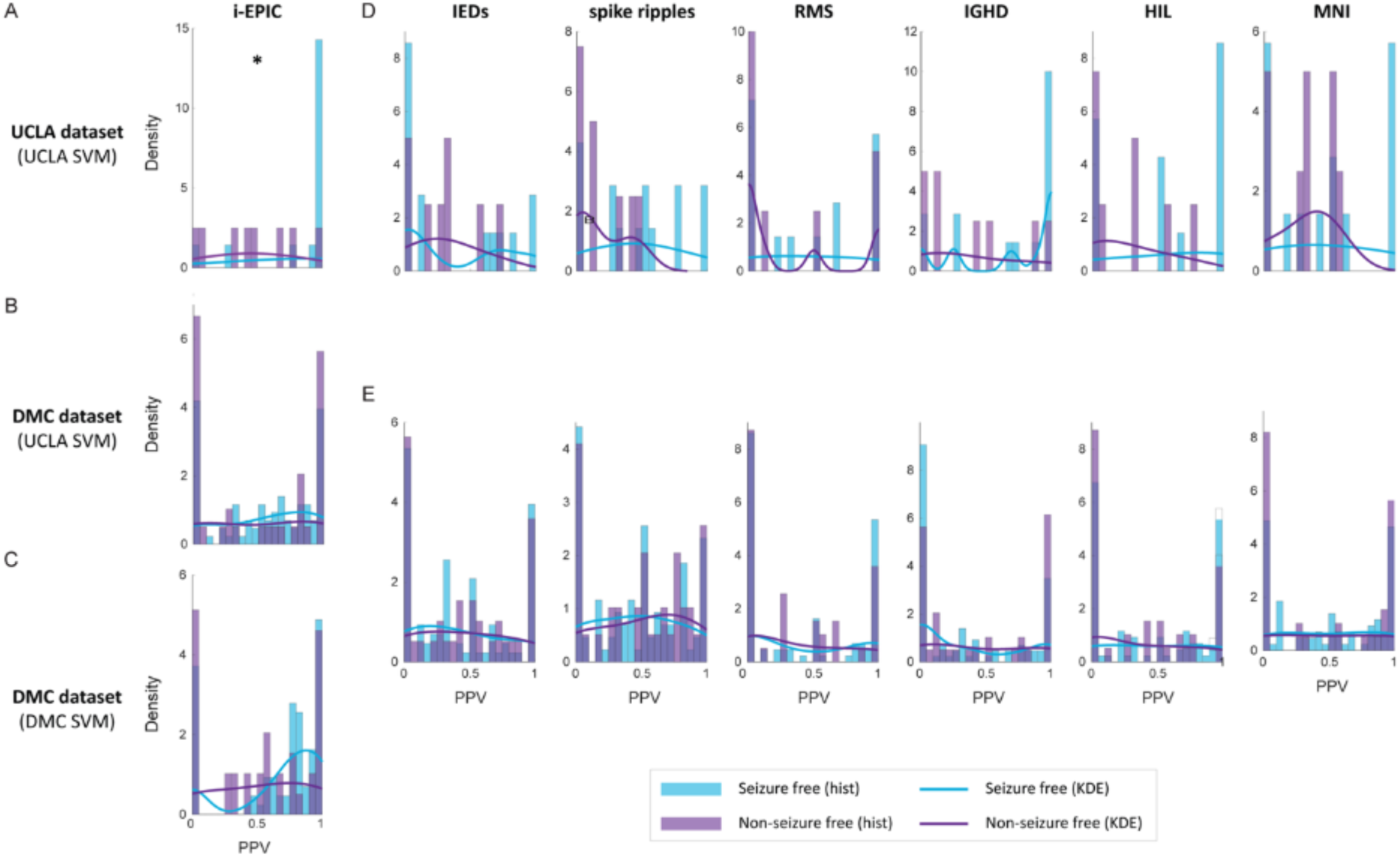
Histograms of PPV values for seizure-free (blue) and non-seizure-free (purple) subjects for the (A, D) UCLA and (B, C, E) DMC datasets. I-EPIC DMC results are shown for two SVM models – (B) trained using UCLA seizure-free subjects, and (C) retraining the SVM with DMC seizure-free data. Each column shows the results from a different algorithm – (A-C) i-EPIC, (D, E) IEDs, spike-ripples, RMS, IGHD, HIL, and MNI detectors (from left to right). Solid lines show the distribution of the histogram values.

In contrast, none of the six benchmark detectors showed statistically significant differences between seizure-free and non-seizure-free subjects in either dataset (Figures 6D-E, Supplementary Figures 3D-E, 4D-E, Wilcoxon rank-sum test). This suggests i-EPIC has greater potential to improve discrimination of epileptogenic tissue for surgical planning.

## 4. Discussion

This study introduces i-EPIC (Intracranial EEG Pattern Identification and Categorization), a novel unsupervised machine learning tool that enables automated biomarker discovery. Our application of i-EPIC to two datasets revealed candidate biomarkers spanning both low (1-51 Hz) and high (70-210 Hz) frequency ranges that can: (1) identify epileptogenic tissue with superior discriminatory power compared to established markers, (2) maintain consistent performance across independent datasets, and (3) potentially aid in prediction of surgical outcome. Despite appearing as clear islands in the time-frequency domain, epileptologists reported that many of the events identified by i-EPIC were not visually discernable in the iEEG. Together, these results highlight the limitations of traditional, visually defined biomarkers^34^ and motivate the use of data-driven methods for identifying clinically relevant electrophysiological events.

The identification of candidate biomarkers in both low and high frequency ranges affirms the need to consider a broad spectrum of electrophysiological activity when delineating the SOZ. Although HFOs have received substantial attention in recent years, established low-frequency biomarkers such as IEDs are also known to carry clinically relevant information. Rather than prioritizing any single frequency band, our results demonstrate that events spanning a wide range of frequencies can provide strong discriminatory power. Notably, low-frequency events (1-51 Hz) showed classification performance comparable to ripple-band activity. These findings align with recent studies suggesting that low-frequency activity can be equally informative for SOZ identification^50^ and support the inclusion of multiple frequency bands when identifying SOZ biomarkers.

Visual inspection of the events identified with i-EPIC highlighted the added value of the TF analysis. Most events were not clearly discernible in the iEEG time series but were clearly visible in the TF domain, including high-frequency activity, gamma oscillations, and low-amplitude beta bursts. This underscores the advantage of data-driven approaches like i-EPIC in detecting subtle or otherwise hidden electrophysiological patterns that may be missed during standard visual review. Although no cluster’s average TF image resembles the morphology of a spike, IEDs are not absent from the identified events. Averaging TF representations reduces the variable high-frequency spike tail, while preserving the more stable low-frequency components. In addition, variability in spike features led to spike-containing events being distributed across multiple clusters.

The substantial inter-subject variability in biomarker effectiveness observed in our study reflects the complex nature of epilepsy, highlighting the limitations of relying on a single biomarker across patients. Although some individual i-EPIC categories achieved performance comparable to the SVM-based combination in both UCLA and DMC datasets, no single category outperformed the combined model. Further, the specific categories that performed well varied across subjects. As a result, integrating multiple biomarker categories using SVM classification provides more robust and generalizable approach, particularly for application to new subjects or data from different centers. This finding is consistent with previous studies showing improved SOZ localization through multi-biomarker combinations.^51^

The robust performance of i-EPIC on the independent DMC dataset demonstrates the method’s ability to generalize across different centers, recording setups, and sampling frequencies. This is critical, as many proposed biomarker detection methods show reduced or inconsistent performance when applied to data outside the conditions under which they were developed.^6^ Prior work on HFO detection has shown that detector performance depends strongly on dataset-specific parameter optimization, selected frequency ranges, and electrode characteristics, and that applying detectors to new datasets without re-optimizing can lead to substantial performance degradation relative to results reported in the original study.^52^ The consistent performance of i-EPIC on lower sampling rate data (1000 Hz) also suggests broader applicability to clinical settings with more limited recording capabilities, addressing a key practical concern in this field.^50^

The biomarker performance for seizure-free and non-seizure-free subjects varied between datasets, possibly reflecting different surgical approaches and their impact on biomarker validation. In the UCLA dataset, biomarkers showed expected superior performance in seizure-free subjects, where all the epileptogenic tissue had been removed during surgery, compared to non-seizure-free subjects where residual epileptogenic tissue may remain unresected and be misclassified by our metric. However, the DMC dataset only showed borderline significant differences between group performances for PPV, potentially due to more extensive resection strategies (29.2% vs. 22.5% total resected channels; 23.2% vs. 17.5% non-SOZ channels resected for DMC and UCLA, respectively), which may have reduced the perceived difference between epileptogenic and non-epileptogenic tissue.

While i-EPIC shows strong performance, there remain several limitations that should be addressed and explored in future studies. First, the number of seizure-free subjects used for training was relatively small, which may limit the generalizability of some individual biomarker categories. Second, there was substantial inter-subject variability in the proportion of resected channels, which could have affected biomarker performance metrics. Additionally, because resected channels in seizure-free subjects were used as a proxy for epileptogenic tissue, electrode coverage may introduce bias, including the resection of an unknown number of normal channels. Third, due to the limited numbers of subjects, we were unable to evaluate whether biomarker performance varies with underlying pathology. Finally, because i-EPIC is fully unsupervised, the physiological mechanisms underlying many of the candidate biomarkers are unclear, as they do not necessarily share characteristics with known epileptiform waveforms.

Future studies should focus on expanding the training set to include larger, multi-center cohorts, which would improve the robustness and generalizability of i-EPIC across diverse recording conditions and patient populations. Additionally, examining how i-EPIC biomarkers perform across different epilepsy pathologies and integrating imaging data could enhance localization accuracy.

In conclusion, i-EPIC represents a promising new approach for automated, unbiased identification of electrophysiological biomarkers in epilepsy. By including multiple frequency bands and generalizing across datasets, it addresses key limitations of existing methods. Further development and validation of this approach could advance our ability to accurately delineate epileptogenic tissue and improve surgical outcomes for patients with drug-resistant epilepsy.

## Data Availability

The study uses ONLY openly available human data that were originally located at: https://openneuro.org/datasets/ds005398/versions/1.1.1

## Acknowledgements

The authors would like to acknowledge the EEG technologists and other hospital staff, as well as the patients and their families for their selfless contributions to this work.

## Author Contributions

Blanca Romero Milà contributed to study conception and design, development of the computational methods, data analysis, and manuscript writing. Nathan Phi Hoang and Marco Pinto-Orellana contributed to method development and computational implementation. Atsuro Daida, Sotaro Kanai, and Naoto Kuroda contributed to data collection and data interpretation. Shaun A. Hussain, Daniel W. Shrey, Eishi Asano, and Hiroki Nariai provided clinical expertise, computational guidance, and interpretation of the results. Beth A. Lopour supervised the study, contributed to study conception, design, and interpretation of findings, and supervised manuscript preparation. All authors reviewed and approved the final manuscript.

## Data availability

The data used in this study correspond to a subset of an iEEG dataset publicly available on OpenNeuro, available at https://openneuro.org/datasets/ds005398/versions/1.0.1. For this analysis, additional channel information was incorporated to perform bipolar re-referencing of the data. We are currently working on adding the bipolar-referenced signals to the original OpenNeuro dataset, so the data will be publicly available prior to publication of this manuscript.

