## Supplementary Material for "Discovering Novel intracranial EEG Biomarkers of Seizure Generating Tissue through Time-Frequency Analysis"

### Supplementary Methods

#### S1. Data pre-processing

All iEEG data underwent bipolar re-referencing to enhance spatial specificity (Figure 1A). Then we implemented an automated artifact detection algorithm based on the signal envelogram method described by Nejedly et al. [1]. The envelogram is the squared magnitude of the Hilbert-transformed iEEG, standardized by converting to a z-score for each channel. Specifically, we computed envelograms from bandpass-filtered signals in four frequency ranges: 1-51 Hz, 70-210 Hz, 211-350 Hz, and 351-500 Hz. These frequency bands were selected to match the time-frequency decomposition in Section 2.2.2.

Segments of iEEG in which the envelogram exceeded a z-score of 8 in at least two frequency bands simultaneously were marked as artifactual. This threshold was selected to ensure high specificity in artifact detection while preserving potential events of interest. The requirement for artifact presence in multiple frequency bands was based on the characteristic broadband nature of many artifacts, such as sharp transients and DC shifts.

#### S2. Time-frequency decomposition and identification of Events of Interest

The Stockwell transform (S-transform) is a generalization of the short-time Fourier transform and a modification of the continuous wavelet transform, combining advantages from both approaches [2-4]:

$$S(\tau, f) = \int_{-\infty}^{\infty} x(t) \frac{|f|}{\sqrt{2\pi}} e^{\frac{-f^2(\tau-t)^2}{2}} e^{-j2\pi f t} dt \qquad (S1)$$

$S(\tau, f)$ was calculated for all iEEG data from each channel and then whitened using the z-score $H_0$ method. This method was selected due to its robustness against non-stationary background activity and because it enables reconstruction of the whitened signal in the time domain, which is advantageous for clinical interpretation. We then defined the whitened time-frequency decomposition as $X_{t,f}$ where $X$ is the power, $t$ is time, and $f$ is the frequency at which the iEEG was measured. Events of interest were identified by thresholding the whitened time-frequency decomposition $X_{t,f}$. Specifically, the set of all time-frequency points exceeding threshold $P$ was defined as:

$$TF = \{(t,f) | X_{t,f} > P\} \qquad (S2)$$

For each iEEG electrode, high-power time–frequency points were identified in the time–frequency decomposition using a fixed power threshold (P). This threshold was determined using an elbow method that optimized the relationship between the threshold value and the number of detected events, and the resulting value was held constant across all subjects, channels, and frequencies.

High-power points were grouped into discrete time–frequency "islands," defined as connected components in the time–frequency space. Each island was treated as a time–frequency event of interest (TFI/EOI). A minimum duration of 10 ms was imposed to ensure inclusion of oscillatory activity, corresponding to at least five cycles at the maximum analyzed frequency (500 Hz).

Simultaneous or overlapping activity appearing as separate islands in different frequency bands was treated as distinct events, allowing independent characterization of activity across frequencies.

### S3. Event features

As described in Section 2.2.2, the time-frequency decomposition was performed using variable frequency steps: 5 Hz increments for low frequencies (1-51 Hz) and 10 Hz increments for high frequencies (70-500 Hz). Additionally, the two datasets have different sampling frequencies (2000 Hz for UCLA and 1000 Hz for DMC). Therefore, we define the area $a$ of each pixel in the time-frequency decomposition using both the frequency step size and the temporal resolution determined by the sampling frequency:

$$a_{t,f} = \Delta f \; x \; \Delta t = \Delta f \; x \; \frac{1}{fs} \qquad (S3)$$

where $\Delta f$ is the frequency step size (5 Hz or 10 Hz), $\Delta t$ is the temporal resolution, and $fs$ is the sampling frequency. This normalization ensures consistent area calculations across different frequency bands and datasets.

The features are shown in Figure 2 and were calculated as follows:

1. Maximum Power ($MP$): The maximum power within the area of each $TFI$:

$$MP_i = max(\{X_{t,f,} \mid (t,f) \in TFI_i\}) \qquad (S4)$$

2. Frequency of Peak Power ($FPP$): The frequency at which the $TFI$ exhibits maximum power:

$$FPP_i = argmax_f(\{X_{t,f,} \mid (t,f) \in TFI_i\}) \qquad (S5)$$

3. Area ($A$): The sum of the areas of all pixels in the time-frequency decomposition comprising the TFI:

$$A_i = \sum_{(t,f)\in TFI_i} a_{t,f} \qquad (S6)$$

4. Height ($H$): The total frequency bandwidth spanned by the $TFI$, calculated as the sum of the frequency step sizes of all frequency bins contained in the event:

$$H_i = \sum_{f\in TFI_i} \Delta f(f) \qquad (S7)$$

   Where $\Delta f(f)$ is the frequency step size at frequency $f$ (5 Hz for low frequencies, and 10 Hz for high frequencies), and the summation is over all unique frequency bins in $TFI_i$.

5. Length ($L$): The temporal duration of the event:

$$L_i = \sum_{t\in TFI_i} \Delta t \qquad (S8)$$

   Where $\Delta t = \frac{1}{fs}$ and the summation is over all unique time bins in $TFI_i$.

6. Density ($D$): The normalized area of the event in time-frequency space, calculated as the ratio of the event area to the maximum possible area (duration times frequency bandwidth):

$$D_{i,} = \frac{A_{i,}}{L_i * H_i} \qquad (S9)$$

7. Density of Simultaneous Events ($Dsim$): The area of all TF activity above threshold occurring between the start ($T_1$) and end ($T_2$) of the $TFI_i$, normalized by the area of that window not occupied by the $TFI_i$:

$$T_1 = min_t(\{TFI_i\}) \qquad (S10)$$

$$T_2 = max_t(\{TFI_i\}) \qquad (S11)$$

$$Dsim_i = \frac{\sum_{(t,f)\in S_i} a_i}{L_i H_{max} - A_i} \qquad (S12)$$

Where $S_i = \{(t,f) | X_{t,f} > P, T_1 \leq t \leq T_2, (t,f) \notin TFI_i\}$ is the set of time-frequency points above threshold $P$ within the event's temporal window but not part of the event itself. $H_{max}$ is the total height of the time-frequency decomposition, which has a constant value of 500 Hz for the UCLA dataset and 350 Hz for DMC. This feature helps distinguish between isolated events, e.g., an HFO on a quiet background, versus simultaneously occurring events, e.g., an HFO occurring with a spike.

8. Density of Surrounding Events ($Dsur$): The normalized area of events occurring within buffer windows immediately before and after the EOI. This metric quantifies the level of background activity surrounding the event, helping distinguish events occurring in active versus quiet backgrounds:

$$Dsur_i = \frac{\sum_{(t,f)\in B_i} a_i}{2\tau \ x \ H_{max}} \qquad (S13)$$

Where $B_i = \{(t,f) | X_{t,f} > P, t \in [T_1 - \tau, T1] \cup [T_2, T_2 + \tau]\}$ is the set of time-frequency points above threshold $P$ in the buffer regions, and $\tau = 100ms$ defines the buffer windows immediately before and after the EOI.

### S4. Comparison of i-EPIC biomarkers to state-of-the-art biomarkers of epileptogenicity

To detect IEDs in the iEEG, we used a previously published method that quantifies spike morphology through analysis of time-series properties: upslope, instantaneous energy, and downslope [5]. Including the waveform shape and energy for event detection has shown to improve performance [6, 7].

Spike ripples are events in the EEG recordings where an IED occurs simultaneously with an HFO. They have been shown to outperform IEDs in identifying the epileptogenic zone [8]. Therefore, we also compared the classification accuracy to that of a spike ripple detector. For this analysis, we utilized *PyHFO* [9], a Python-based HFO detection and classification tool. Specifically, we employed the MNI detector within *PyHFO*, which integrates deep learning models to classify artifacts and identify HFOs co-occurring with spikes [10].

To compare to automated HFO detection, we implemented four previously published algorithms. These included detectors based on the root-mean-square (RMS) amplitude [11], complemented by its corresponding false HFO detection method [12], and the iterative gamma-fit HFO detector (IGHD) [13]. Additionally, we implemented the Hilbert detector (HIL) [14] and the MNI detector [15] using the RIPPLELAB software package for MATLAB [16].

### Supplementary Figures

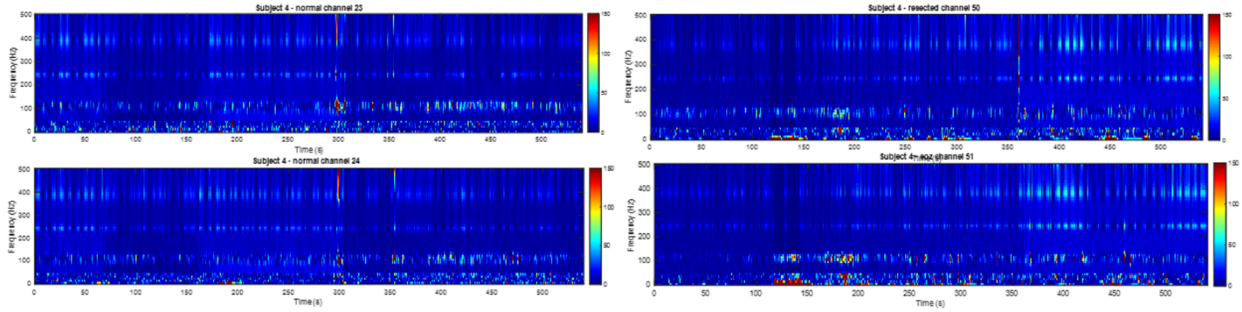


***Supplementary Figure 1:*** *Representative examples of iEEG channels excluded from analysis following visual inspection, due to persistent artifact contamination.*

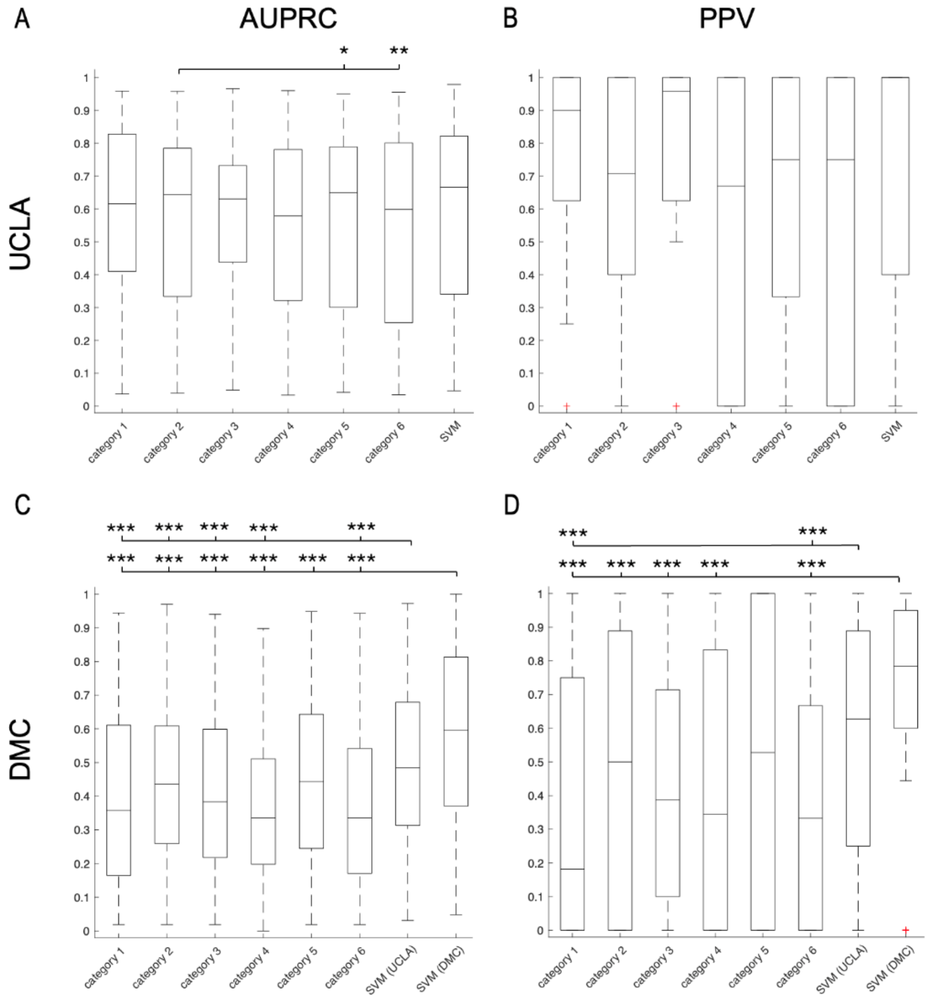


***Supplementary Figure 2:*** *Boxplots illustrate the distribution of (A, C) AUPRC and (B, D) PPV for the six EOI categories present in all subjects compared to i-EPIC leave-one-out cross-validation. Results are shown for the seizure-free patients in the (A, B) UCLA and (C, D) DMC datasets. Central lines represent medians, boxes show interquartile ranges, and whiskers extend to the most extreme non-outlier data points. Statistical significance determined by Wilcoxon signed rank test and BH adjustment (*: p<0.05; **: p<0.01; ***: p<0.001).*

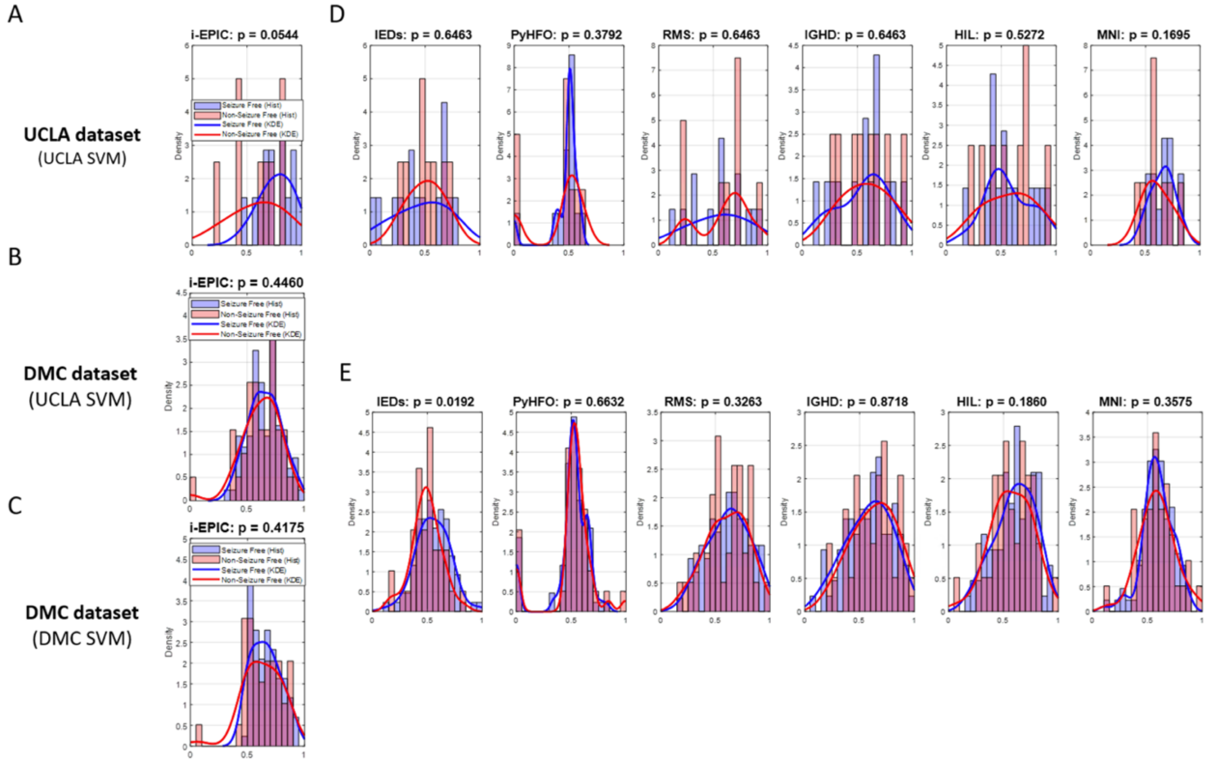


***Supplementary Figure 3:*** *Histograms of the distribution of AUROC values between seizure-free (blue) and non-seizure-free (red) subjects for the (A, D) UCLA and (B, C, E) DMC datasets. I-EPIC DMC results are shown for two SVM models – (B) trained using UCLA seizure-free subjects, and (C) retraining the SVM with DMC seizure-free data. Each column shows the results from a different algorithm – (A-C) i-EPIC, (D, E) IEDs, spike-ripples, RMS, IGHD, HIL, and MNI detectors (from left to right). Solid lines show the distribution of the histogram values.*

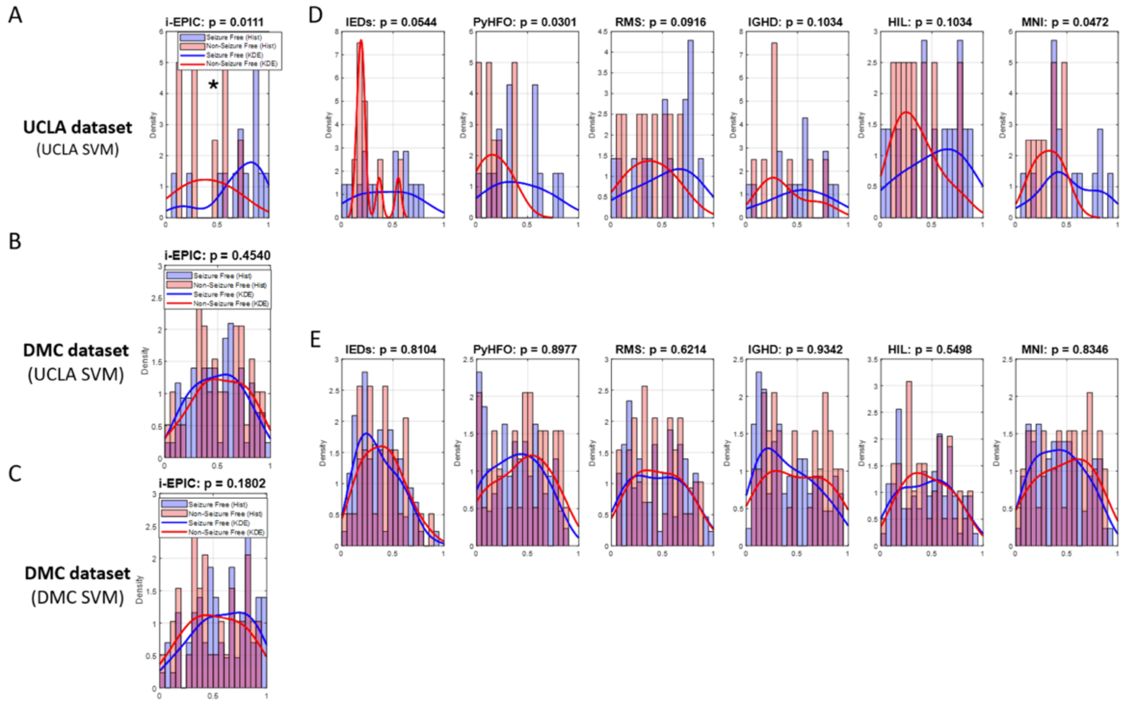


***Supplementary Figure 4:*** *Histograms of the distribution of AUPRC values between seizure-free (blue) and non-seizure-free (red) subjects for the (A, D) UCLA and (B, C, E) DMC datasets. I-EPIC DMC results are shown for two SVM models – (B) trained using UCLA seizure-free subjects, and (C) retraining the SVM with DMC seizure-free data. Each column shows the results from a different algorithm – (A-C) i-EPIC, (D, E) IEDs, spike-ripples, RMS, IGHD, HIL, and MNI detectors (from left to right). Solid lines show the distribution of the histogram values.*

| | UCLA dataset | | | | | | | | DMC dataset | | | | | | | |
|---|---|---|---|---|---|---|---|---|---|---|---|---|---|---|---|---|
| | FPP | MP | Area | Length | Height | Density | Dsim | Dsur | FPP | MP | Area | Length | Height | Density | Dsim | Dsur |
| 1 | 1 - 51 | high | high | high | high | low | high | high | 1 - 51 | high | high | high | high | low | high | high |
| 2 | 1 - 51 | high | high | low | high | low | high | high | 1 - 51 | high | high | low | high | low | high | high |
| 3 | 1 - 51 | high | high | low | high | low | high | low | 1 - 51 | high | high | high | high | low | high | low |
| 4 | 1 - 51 | high | high | high | high | low | high | low | 1 - 51 | low | low | low | high | low | high | high |
| 5 | 1 - 51 | high | low | low | high | low | high | high | 70 - 210 | high | high | high | high | low | high | low |
| 6 | 1 - 51 | low | low | low | high | low | high | high | 1 - 51 | low | low | low | high | high | high | high |
| 7 | 70 - 210 | high | high | high | high | low | high | low | 70 - 210 | high | high | high | high | low | high | high |
| 8 | 1 - 51 | low | low | low | high | high | high | high | 70 - 210 | high | high | low | high | low | high | high |
| 9 | 70 - 210 | low | low | low | low | high | high | high | 70 - 210 | high | high | high | low | low | high | high |

***Supplementary Table 1:*** *9 top performing event categories when developing i-EPIC using UCLA and DMC seizure-free data. Categories highlighted using color are common to both datasets, while categories with a white background appear in only one of the datasets.*

15. Zelmann R, Mari F, Jacobs J, Zijlmans M, Dubeau F, Gotman J. A comparison between detectors of high frequency oscillations. 2012 123(1):106-16. https://www.sciencedirect.com/science/article/pii/S1388245711004263.
16. Navarrete M, Alvarado-Rojas C, Le Van Quyen M, Valderrama M. RIPPLELAB: A Comprehensive Application for the Detection, Analysis and Classification of High Frequency Oscillations in Electroencephalographic Signals. PLoS ONE. 2016 11.
